# Normalizing Spinal Cord Compression Morphometric Measures: Application in Degenerative Cervical Myelopathy

**DOI:** 10.1101/2024.03.13.24304177

**Authors:** Sandrine Bédard, Jan Valošek, Maryam Seif, Armin Curt, Simon Schading, Nikolai Pfender, Patrick Freund, Markus Hupp, Julien Cohen-Adad

## Abstract

**Objective:** Automatic and robust characterization of spinal cord shape from MRI images is relevant to assess the severity of spinal cord compression in degenerative cervical myelopathy (DCM) and to guide therapeutic strategy. Despite its popularity, the maximum spinal cord compression (MSCC) index has practical limitations to objectively assess the severity of cord compression. Firstly, it is computed by normalizing the anteroposterior cord diameter by that above and below the level of compression, but it does not account for the fact that the spinal cord itself varies in size along the superior-inferior axis, making this MSCC sensitive to the level of compression. Secondly, spinal cord shape varies across individuals, making MSCC also sensitive to the size and shape of every individual. Thirdly, MSCC is typically computed by the expert-rater on a single sagittal slice, which is time-consuming and prone to inter-rater variability. In this study, we propose a fully automatic pipeline to compute MSCC.

**Methods:** We extended the traditional MSCC (based on the anteroposterior diameter) to other shape metrics (transverse diameter, area, eccentricity, and solidity), and proposed a normalization strategy using a database of healthy adults (n=203) to address the variability of the spinal cord anatomy between individuals. We validated the proposed method in a cohort of DCM patients (n=120) with manually derived morphometric measures and predicted the therapeutic decision (operative/conservative) using a stepwise binary logistic regression including demographics, clinical scores, and electrophysiological assessment.

**Results:** The automatic and normalized MSCC measures significantly correlated with clinical scores and predicted the therapeutic decision with higher accuracy than the manual MSCC. Results show that the sensory dysfunction of the upper extremities (mJOA subscore), the presence of myelopathy and the proposed MRI-based normalized morphometric measures were significant predictors of the therapeutic decision. The model yielded an area under the curve of the receiver operating characteristic of 80%.

**Conclusion:** The study introduced an automatic method for computation of normalized MSCC measures of cord compression from MRI scans, which is an important step towards better informed therapeutic decisions in DCM patients. The method is open-source and available in the Spinal Cord Toolbox v6.0.

## 1. Introduction

Morphometric measures computed from structural magnetic resonance imaging (MRI) scans are often used to evaluate the severity of spinal cord compressions in degenerative cervical myelopathy (DCM)^1–3^. Morphometric measures also demonstrated potential in predicting DCM progression^4,5^ and to guide therapeutic strategy^6–8^. Nonetheless, these measures present considerable variability due to anatomical differences between spinal levels and among individuals, currently limiting their usage.

Variations in spinal cord anatomy along the superior-inferior direction within a single individual lead to different morphometric measures between levels. For example, spinal cord measures at the C2 vertebral level differ from those at the C4 vertebral level^9–11^. Maximum spinal cord compression (MSCC) is a popular index providing compression severity normalization by using the non-compressed levels above and below the compression site^12,13^. However, MSCC does not consider the varying spinal cord anatomy across spinal levels, which can lead to inaccuracies in the assessment of compression severity, as the measures may not reflect the true pathology extent. For instance, if a compression occurs at the cervical enlargement (around C5 level), the MSCC^13^ would be underestimated because the levels above and below the compression site are typically smaller than the cervical enlargement. Analogously, if the compression happens right below the cervical enlargement, the opposite would happen: the MSCC would be overestimated. Additionally, the MSCC computation is usually done manually by clinicians on a single sagittal slice which is time-consuming and prone to variability between experts.

Inter-subject variability, on the other hand, is associated with the differences between individuals in terms of age, sex, and body size (weight, height). For instance, a significantly smaller spinal cord area is consistently reported in females relative to males^11,14,15^. Similarly, age-related changes in spinal cord size and shape can also impact spinal cord morphometrics and may require different normalization strategies for different age groups^15^. Previous studies proposed different normalization strategies to mitigate this variability, like normalizing to age-matched healthy control^16^ or taking into account confounding variables like age and brain volume^14,15,17^. However, an age– and sex-matched healthy control cohort is not always available, measures like brain volume are not commonly accessible within spinal cord studies, and there is no clear consensus on which demographic and anatomical factors to use for normalization.

In this study, we aimed to improve the accuracy of spinal cord compression morphometric measures by mitigating the effects of intra-subject and inter-subject variability. We proposed an automatic computation of the MSCC from MRI scans and extended it to other morphometric measures: transverse (RL) diameter, cross-sectional area (CSA), eccentricity, and solidity. Furthermore, we implemented an inter-subject normalization to account for the varying anatomy along the spinal cord. We validated our approach in a large cohort of DCM patients (n=120). The proposed method is available as part of the open-source software Spinal Cord Toolbox (SCT).

## 2. Materials and methods

In this section, we first describe the automatic self-normalization of morphometric measures using the non-compressed levels above and below the compression site. Then, we introduce the normalization across subjects based on a database of healthy adults. Finally, we validate the method in a cohort of DCM patients.

### 2.1. Self-normalizing of morphometric measures

MSCC is typically used as a self-normalized metric to assess compression severity computed by measuring the anteroposterior (AP) diameter at the level of compression and at the levels above and below the compression site^13^. The computation is usually done manually by an expert-rater on a single sagittal slice which is a time-consuming process prone to inter-rater variability.

In the current study, we automated the MSCC computation and included additional metrics besides the AP diameter (<u>2.1.1</u> and <u>2.1.2</u>). We refer to these metrics as *morphometric ratios*. Additionally, we added an across-subject normalization to consider the varying spinal cord anatomy along the superior-inferior axis between subjects (<u>2.1.3</u>). The method is available in SCT’s v6.0 and higher via the `sct_compute_compression` function.

#### 2.1.1. Automatic MSCC computation

Figure 1 presents an overview of the automatic morphometric ratios computation. Given that compression typically affects more than one axial slice, and compression sites are commonly located near the intervertebral discs, we opted not to directly take the entire vertebral levels above and below the compression site for self-normalization. Instead, we averaged the metrics on 20mm extent at a 10mm distance above and below the compression site. If multiple compressions are present, we use 10mm above the most upper compression, and 10mm below the lowest compression site to ensure that “non-compressed” levels are used for the normalization. We chose an extent of 20mm to approximate vertebral level length^10^, and a distance of 10mm to reliably select only non-compressed areas. The extent and distance can be specified by the user as input arguments of the `sct_compute_compression` function.

**Figure 1.**
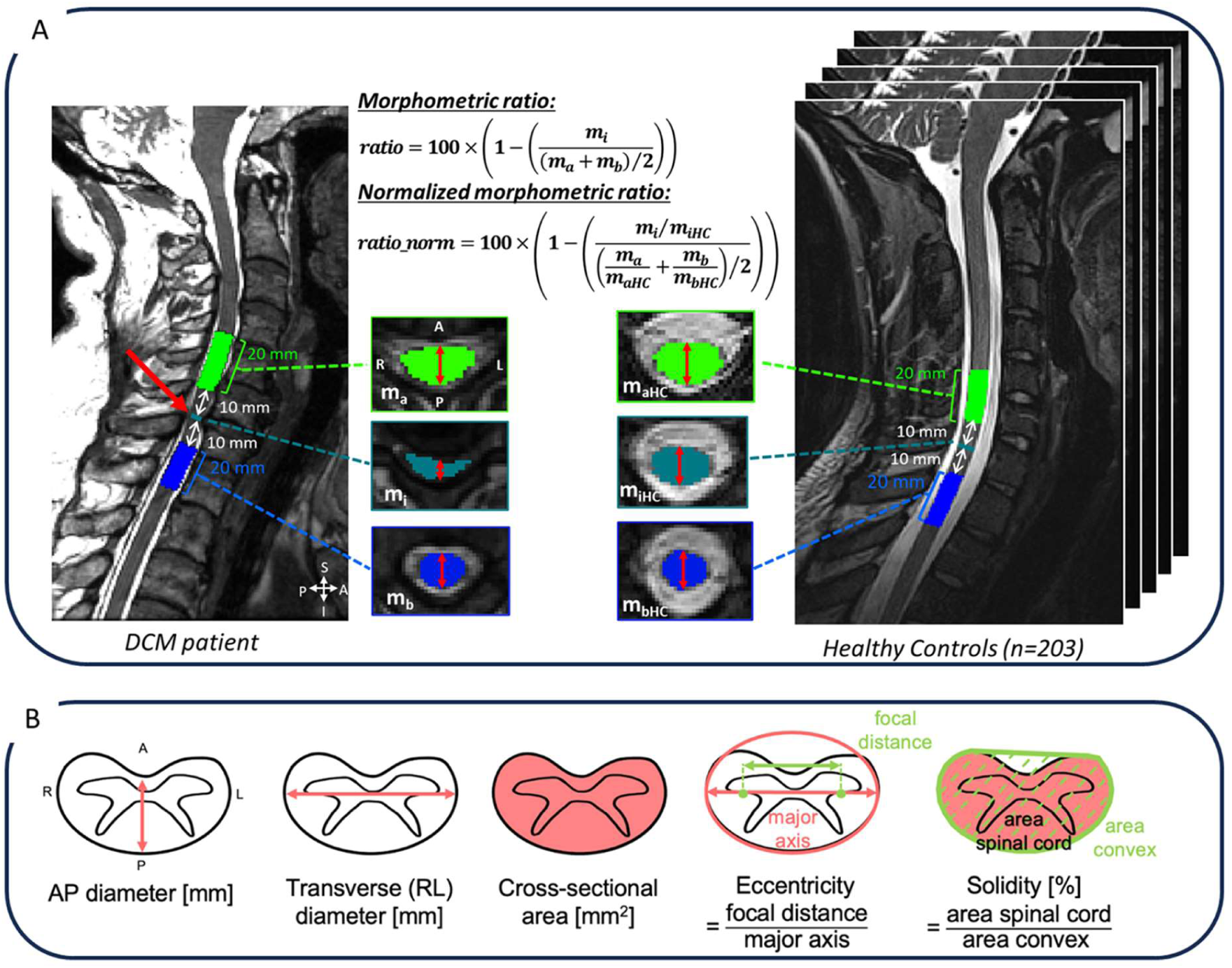
(**A**) Morphometric ratio and normalized morphometric ratio. The morphometric ratio is computed based on the metric (here AP diameter is shown) at the compression site, 10mm above and below the compression and averaged along a 20mm extent on the patient and healthy controls. Normalized morphometric ratio (ratio_norm) is computed by dividing each metric (m_x_) by the corresponding value averaged across healthy controls. **(B)** Morphometric measures used to compute morphometric ratios and normalized morphometric ratios. Adapted from^11^.

#### 2.1.2. Morphometric measures to characterize spinal compression

Various morphometric measures were shown to be sensitive to spinal cord compression^16,17^ and DCM progression^4,5^. Therefore, we implemented the normalization approach for several morphometric measures provided by SCT’s sct_process_segmentation. Those metrics are: 1) AP diameter, 2) RL diameter, 3) CSA, 4) eccentricity, and 5) solidity^11^. Note that the original MSCC normalizes only the AP diameter^13^ (see Figure 1B).

The sct_process_segmentation requires a spinal cord segmentation mask, vertebral labels, and a file with identified compression sites as inputs and outputs a CSV file with computed metrics for each compression site. A tutorial on how to get the labels and use sct_process_segmentation is available at: https://spinalcordtoolbox.com/user_section/tutorials/shape-analysis/normalize-morphometrics-compression.html.

#### 2.1.3. Normalization with healthy controls

To account for variability caused by anatomical differences between spinal levels, the self-normalized morphometrics were additionally computed on a database of healthy adult volunteers (n=203) in the PAM50 anatomical dimensions (Figure 1A)^11^. To account for the influence of sex and age on the morphometric measures, we implemented an option to filter healthy controls based on sex (males vs females) and a specific age range (e.g., 40-50 y.o.).

### 2.2. Validation in patients with cord compression

In the following section, we validated the automatic morphometric ratios against manually derived shape measures in a single-center DCM cohort. We also explored the relationship of the morphometric ratios with clinical scores and examined which metrics can predict the therapeutic decision (operative/conservative).

#### 2.2.1. Participants

120 DCM patients were recruited at Balgrist University Hospital (Zurich, Switzerland) between October 2016 and December 2022. The research received approval from the regional ethical review board (Kantonale Ethikkommission Zurich, KEK-ZH 2012-0343, BASEC Nr. PB_2016-00623), and is registered (www.clinicaltrials.gov). The execution of the study adhered to the ethical guidelines set forth by the World Medical Association’s Declaration of Helsinki, which pertains to human experimentation. Prior to their inclusion in the study, all participants gave their informed consent.

Inclusion criteria: cervical spinal stenosis on the T2-weighted (T2w) MRI; clinical symptoms consistent with DCM^3^ (i.e., pain, sensory or motor deterioration in the upper or lower limbs, gait or bladder dysfunction); age 18-80 years. Patients suffering from a competing neurological disease with a potential bias to clinical and neurophysiological assessments were excluded. Additional exclusion criteria: MRI contraindications, epileptic seizures, mental illness, severe medical illness and pregnancy.

#### 2.2.2. Clinical and electrophysiological assessment

Neurological status and functional impairment were assessed with the following scores:

1. Modified Japanese Orthopedic Scale (mJOA, max. 18 points)^18^ and its subscores upper extremity sensory dysfunction, lower extremity motor dysfunction, upper extremity motor dysfunction and bladder dysfunction.
2. American Spinal Injury Association (ASIA)/Graded Redefined Assessment of Strength, Sensibility, and Prehension (GRASSP) subscores^19,20^: the upper-extremity light-touch score, the upper-extremity pin prick score (cervical pin prick score), monofilament testing of dorsal hand sensation (monofilament sensation score), upper extremity motor score and lower extremity motor score.

Electrophysiological assessment included dermatomal somatosensory evoked potentials (dSEP) from spinal levels C6 and C8 and contact heat evoked potentials (CHEPS) from levels C6, C8, and T4. The electrophysiological assessment was performed as previously described in Scheuren et al. ^21^

Moreover, the following clinical information was provided: level of maximal compression, number of stenosis levels, spine surgery prior to study inclusion (0-no, 1 –yes), and presence of radiological signs of myelopathy (i.e., T2w hyperintensity; 0 – no, 1 – yes). The presence of a hyperintense T2w signal (e.g., diffuse T2 hyperintensity, cystic lesions, snake eyes) within the spinal cord (representing radiological signs of established myelopathy) was visually evaluated in axial and sagittal T2w MRI (<u>2.2.3.</u>).

#### 2.2.3. MRI acquisition

MRI scanning was performed on a 3T scanner (Siemens SkyraFit, Erlangen, Germany), using the body transmit coil and the product 16-channel receive head/neck coil. An MRI compatible cervical collar was used to reduce involuntary neck motion. The axial T2w scans were acquired with the following parameters: TE of 93 ms, TR of 3600 ms, slice thickness of 3mm, flip angle of 150 degrees, field-of-view of 160mm, bandwidth of 284 Hz/px, base resolution of 320, phase resolution of 80%, spatial resolution of 0.5×0.5×3.0mm³, and GRAPPA 2. The sagittal T2w scans were were acquired with the following parameters: TE of 87 ms, TR of 3760 ms, slice thickness of 2.5mm, flip angle of 160 degrees, field-of-view of 220mm, bandwidth of 260 Hz/px, base resolution of 384, phase resolution of 75%, spatial resolution of 0.6×0.6×2.5mm³.

#### 2.2.4. MRI analysis (manual)

Clinicians performed a manual assessment of the MRI images using the adapted spinal canal occupation ratio (aSCOR) calculated at the level of compression as the ratio of the spinal cord area divided by the spinal canal area multiplied by 100^22,23^ and adapted MSCC (aMSCC) calculated as the ratio of CSA in the compressed segment divided by the CSA at C2 vertebral level.

#### 2.2.5. MRI analysis (automatic)

Automatic processing was done using SCT v6.0^24^ and the dcm-metric-normalization pipeline (https://github.com/sct-pipeline/dcm-metric-normalization) and is illustrated in Figure 2. For each subject, intervertebral discs were obtained from the sagittal T2w image using sct_label_vertebrae^25^, and the spinal cord was segmented on the axial T2w image using sct_deepseg_sc^26^ (Figure 2A). The intervertebral discs and spinal segmentations were visually controlled and manually corrected when necessary. Then, the sagittal T2w image was resampled to the axial T2w image to bring sagittal intervertebral discs to the axial scan and to label the axial spinal cord segmentation (Figure 2B). The compression sites were manually identified on the axial T2w images based on the clinical information (Figure 2B). Finally, the `sct_compute_compression` function was used to automatically compute morphometric ratios and normalized morphometric ratios (<u>2.1.3</u>) (Figure 2C). To mitigate the differences in morphometrics between sex when computing the normalized morphometric ratios, we filtered healthy controls according to sex. Morphometric ratios and normalized morphometric ratios were computed for five measures (AP diameter, RL diameter, CSA, eccentricity, and solidity).

**Figure 2.**
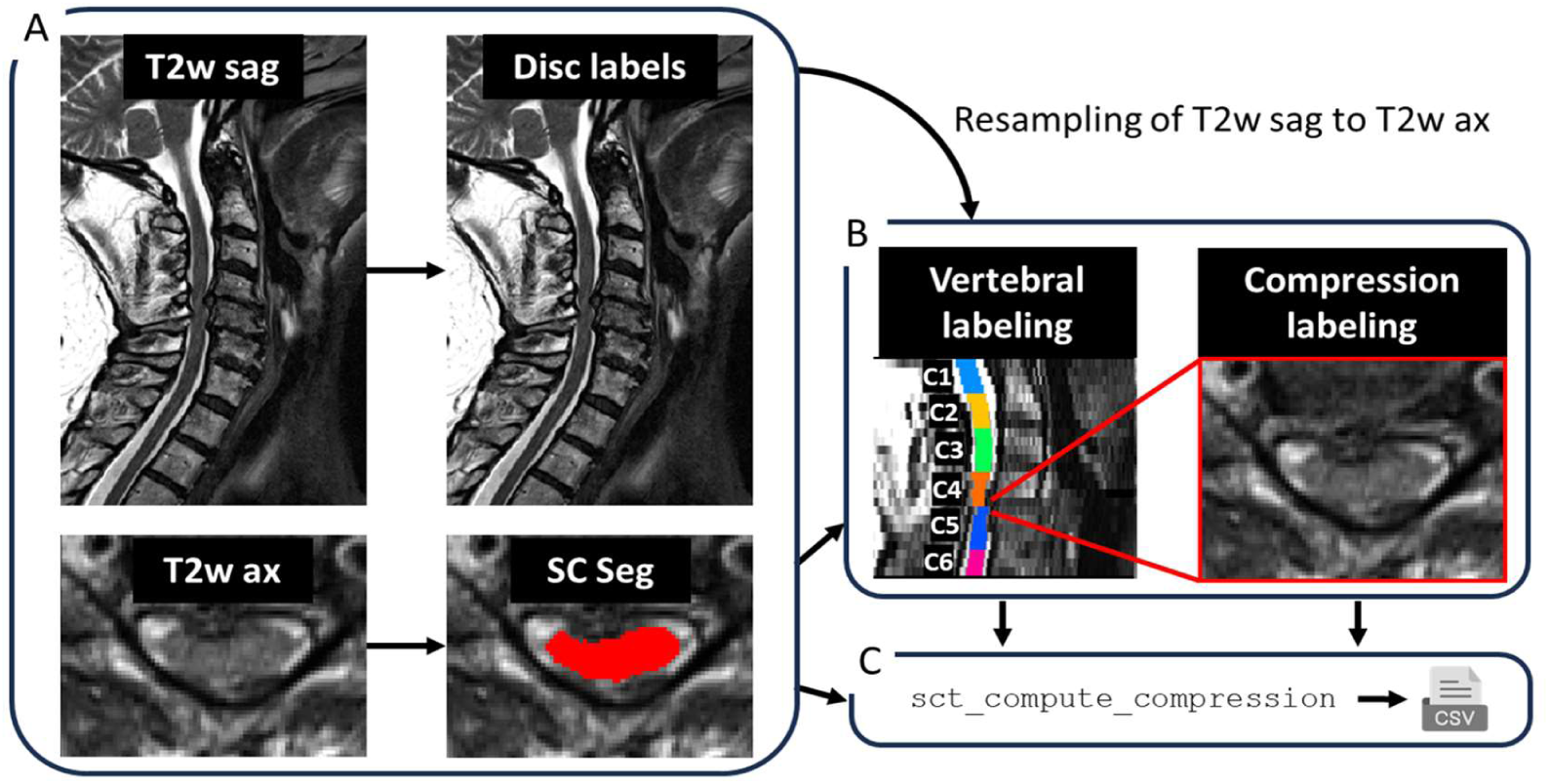
Automatic processing pipeline. Intervertebral discs were obtained for the sagittal T2w image, and the spinal cord was segmented on the axial T2w image (A). Then, the sagittal T2w image was resampled to the axial T2w images to bring the intervertebral discs from sagittal to axial T2w scan to label the axial spinal cord segmentation (B). Spinal cord compression sites were manually labeled, and morphometric ratios and normalized morphometric ratios were automatically computed using the sct_compute_compression function (C).

#### 2.2.6. Statistics

Statistical analysis was performed with SciPy Python library v1.6.3 and scikit-learn Python library v0.23.2. Descriptive statistics including mean and standard deviation were provided. We computed the Spearman correlation among all available regressors (as provided in **Table 1**) between continuous variables. Phi coefficient was computed across dichotomous variables and the point-biserial correlation was computed across continuous and dichotomous variables. Dichotomous variables included sex, previous surgery, myelopathy and therapeutic decision. We did not correct for multiple comparisons when computing correlations since the purpose was exploratory and the number of regressors is considered in the subsequent multivariate regression analysis.

**Table 1.** Descriptive statistics of demographics, clinical data, clinical scores, morphometrics, and electrophysiological data.

| Demographics |  |  |
| --- | --- | --- |
| Age | 55.28 ± 12.80 |  |
| Sex (M: male/F: female) | M: 64% F: 36% |  |
| Height (m) | 1.70 ± 0.10 |  |
| Weight (kg) | 74.93 ± 14.17 |  |
| Clinical data |  |  |
| Previous surgery (yes/ no) | 6% | 94% |
| Therapeutic decision (conservative/operative) | 40% | 60% |
| Myelopathy (yes/no) | 38% | 62% |
| Number of stenoses | 2.09 ± 0.97 |  |
| Maximum level of compression | C3/C4: 17%<br>C4/C5: 23%<br>C5/C6: 52%<br>C6/C7: 8% |  |
| Clinical scores |  |  |
| mJOA (max. 18 points) | 15.89 ± 1.84 |  |
| Upper extremity motor subscore (max 5 points) | 4.54 ± 0.67 |  |
| Lower extremity motor subscore (max 7 points) | 6.49 ± 0.80 |  |
| Upper extremity sensory subscore (max 3 points) | 2.18 ± 0.64 |  |
| Bladder function subscore (max 3 points) | 2.68 ± 0.58 |  |
| Monofilament sensation score (max 24 points) | 21.78 ± 2.80 |  |
| Cervical light touch score (max 28 points) | 25.83 ± 3.16 |  |
| Cervical pin prick score (max 28 points) | 25.16 ± 3.84 |  |
| Upper extremity motor score ASIA (max 50 points) | 49.34 ± 1.40 |  |
| Lower extremity motor score ASIA (max 50 points) | 49.58 ± 1.51 |  |
| Morphometric ratios (automatic, normalized) |  |  |
| AP diameter ratio (%) | 18.51 ± 11.27 |  |
| AP diameter ratio normalized (%) | 17.20 ± 9.96 |  |
| CSA ratio (%) | 18.42 ± 15.36 |  |
| CSA ratio normalized (%) | 20.38 ± 12.81 |  |
| RL diameter ratio(%) | -5.40 ± 12.64 |  |
| RL diameter ratio normalized (%) | 0.45 ± 9.42 |  |
| Eccentricity ratio (%) | -8.29 ± 4.91 |  |
| Eccentricity ratio normalized (%) | -4.93 ± 3.80 |  |
| Solidity ratio (%) | 5.06 ± 5.56 |  |
| Solidity ratio normalized (%) | 4.48 ± 4.98 |  |
| Morphometric ratios (manual) |  |  |
| aSCOR (%) | 75.52 ± 13.16 |  |
| aMSCC (%) | 100.57 ± 17.38 |  |
| Electrophysiological data |  |  |
| dSEP C6 pathologic | n=5 |  |
| dSEP C8 pathologic | n=9 |  |
| CHEPS C6 pathologic | n=42 |  |
| CHEPS C8 pathologic | n=69 |  |
| CHEPS T4 pathologic | n=29 |  |

##### 2.2.6.1. Correlation between manual, automatic and normalized morphometrics

First, we computed the Spearman’s correlation between the manual measures (aSCOR and aMSCC), and automatic morphometric ratios. As aSCOR and aMSCC were computed using the CSA, we only compared them with CSA ratio and CSA ratio normalized

##### 2.2.6.2. Prediction of therapeutic outcome

To validate the automatic computation of morphometric ratios, we created a model to predict the therapeutic decision (conservative/operative) using a stepwise binary logistic regression. The dependent variable was the therapeutic decision (conservative: 0, operative: 1). The independent variables included demographics, clinical data, clinical scores, electrophysiological data, automatic morphometric ratios, and manual morphometric measures as detailed in **Table 1**.

First, we proceeded to a stepwise logistic regression with p-value=0.05 as the level of significance. A binary logistic regression was generated using the resulting significant variables of the stepwise regression. The model was validated using a 10-fold cross-validation. The receiver operating characteristic (ROC) curve and area under the curve (AUC) were used as performance metrics. We repeated the process using the normalized morphometric ratios, leading to 2 different models. If the participant’s maximum level of compression was not included in the axial field-of-view, the participant was excluded from the analysis.

## 3. Results

### 3.1. Descriptive statistics

Detailed demographics and descriptive statistics for all available regressors are provided in **Table 1**. Quantitative variables are represented as mean±standard deviation while categorical variables are presented as % in each category.

### 3.2. Correlations between morphometrics and clinical scores

The correlation matrix across all variables is presented in the supplementary materials **Figure S1**. Some correlations are described in the following.

#### 3.2.1. Correlations with clinical data

The therapeutic decision significantly (p-value<0.05) correlated with the mJOA (ρ=-0.57), upper extremity motor subscore (ρ=-0.45), lower extremity motor subscore (ρ=-0.50), upper extremity sensory subscore (ρ=-0.26), bladder function subscore (ρ=-0.26), radiological signs of myelopathy (ρ=0.38), upper extremity motor score ASIA (ρ=-0.28), lower extremity motor score ASIA (ρ=-0.27), cervical light touch score (ρ=-0.40), the cervical pin prick score, (ρ=-0.39), and the monofilament sensation score (ρ=-0.35).

Among the automatic morphometrics, AP diameter ratio (ρ=0.36), AP diameter ratio normalized (ρ=0.32), CSA ratio (ρ=0.43), CSA ratio normalized (ρ=0.38), solidity ratio (ρ=0.28), and solidity ratio normalized (ρ=0.28) were significantly correlated with the therapeutic decision. Normalized ratios seem to have lower correlations with the therapeutic decision than the ratios without normalization. For the manual morphometrics, aSCOR correlated significantly with the therapeutic decision (ρ=0.24), while aMSCC did not significantly correlate with the therapeutic decision.

Radiological signs of myelopathy significantly correlated with AP diameter ratio (ρ=0.43), AP diameter ratio normalized (ρ=0.41), CSA ratio (ρ=0.55), CSA ratio normalized (ρ=0.53), RL diameter ratio (ρ=0.31), RL diameter ratio normalized (ρ=0.32), and aSCOR (ρ=0.30) while it did not correlate significantly with aMSCC.

The number of stenoses significantly correlated with age (ρ=0.21), potentially suggesting that the number of stenoses increased with age.

#### 3.2.2. Correlations with clinical scores

Correlations between CSA ratio, CSA ratio normalized, AP diameter ratio, AP diameter ratio normalized, RL diameter ratio and RL diameter ratio normalized were significant with motor dysfunction of lower extremities and upper extremities (lower extremity motor subscore: – 0.33<ρ<-0.22, and upper extremity motor subscore: –0.36<ρ<-0.19). The mJOA correlated significantly with AP diameter ratio, CSA ratio and CSA ratio normalized (−0.29<ρ<-0.22).

The upper extremity motor score ASIA correlated with CSA ratio (−0.25, p-value=0.013) and RL diameter ratio (−0.24, p-value=0.015). The lower extremity motor score ASIA correlated with AP diameter ratio (−0.23, p-value=0.024), CSA ratio (−0.31, p-value=0.013), CSA ratio normalized (−0.23, p-value=0.021), and RL diameter ratio (−0.22, p-value=0.03). The cervical light touch score correlated with CSA ratio (ρ=-0.18) and CSA ratio normalized (ρ=-0.18).

aSCOR correlated significantly with lower extremity motor subscore (ρ=-0.28). aMSCC correlated with the cervical pin prick score (ρ=0.2).

### 3.3. Correlation between manual, automatic and normalized morphometrics

Figure 3 presents the Spearman’s correlation and scatterplots between the manual morphometric ratios performed by physicians (aSCOR, aMSCC) and proposed automatic morphometric ratios (CSA ratio and CSA ratio normalized).

**Figure 3.**
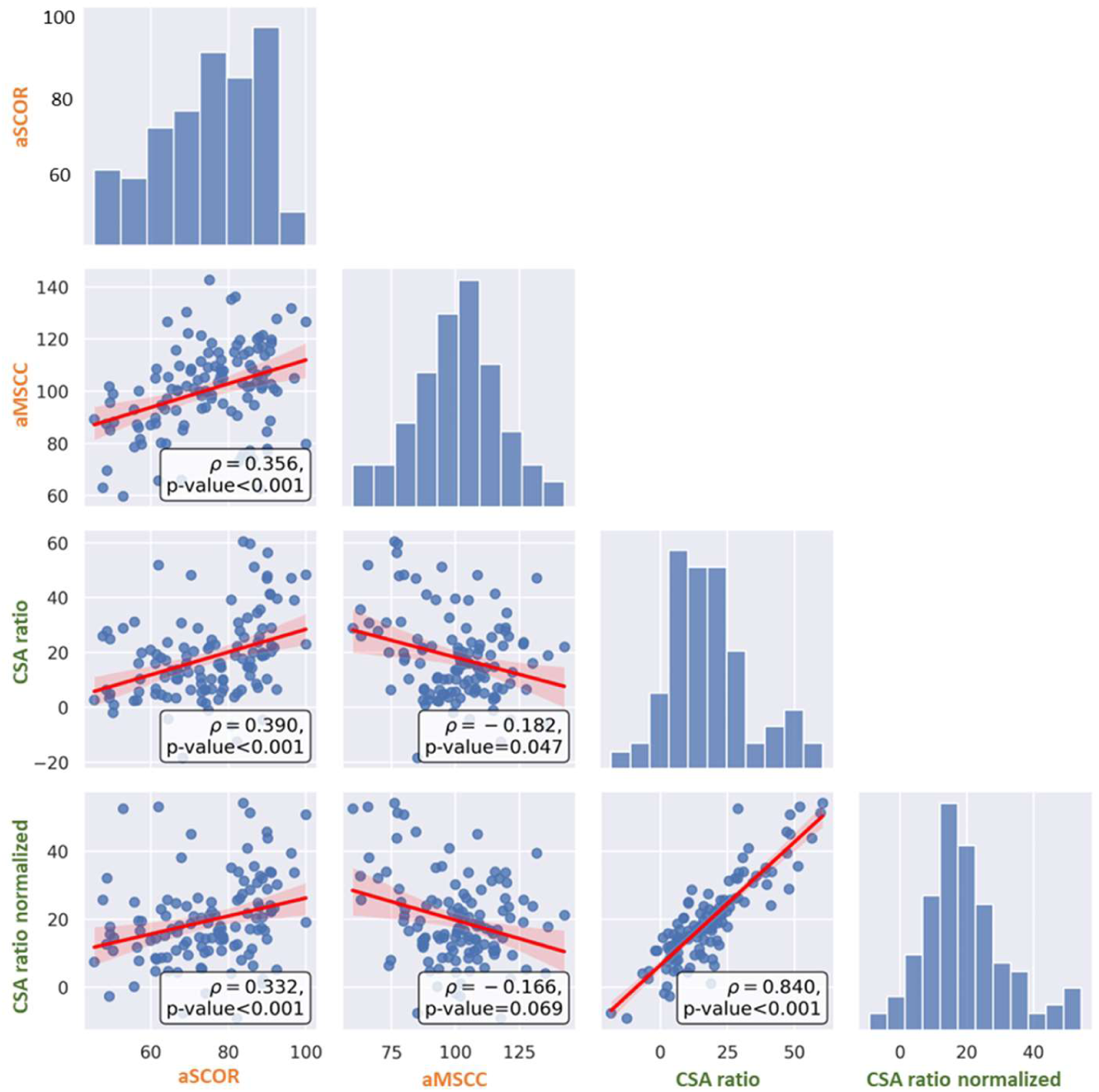
Spearman’s correlation between manual measures (aSCOR, aMSCC) in orange, and automatic morphometric ratios (CSA ratio, CSA ratio normalized) in green.

### 3.4. Predicting therapeutic decision

Using the available regressors described in **Table 1**, we tested if therapeutic decision of the DCM cohort (operative/conservative) could be predicted using a stepwise binary logistic regression. We developed 2 models: (1) without normalization of the automatic morphometric ratios and (2) with normalization of the automatic morphometric ratios. We tested these models in two datasets: one that included all the regressors (n=54 patients), and one without the electrophysiological data (n=100).

#### 3.4.1. Using all the regressors

We first used all the available regressors (**Table 1**) to predict the therapeutic decision. Participants with missing data were excluded (n=46 without electrophysiological data,n=20 without clinical scores), bringing the total number of participants from 120 to 54.

For both models using the morphometrics ratios with and without normalization with healthy controls, the significant regressors are presented in **Table 2** with the resulting logistic regression model. The ROC curves are presented in Figure 4 without normalization (Figure 4a) and with normalization (Figure 4b). Both models yielded an accuracy of 0.767±0.152.

**Figure 4.**
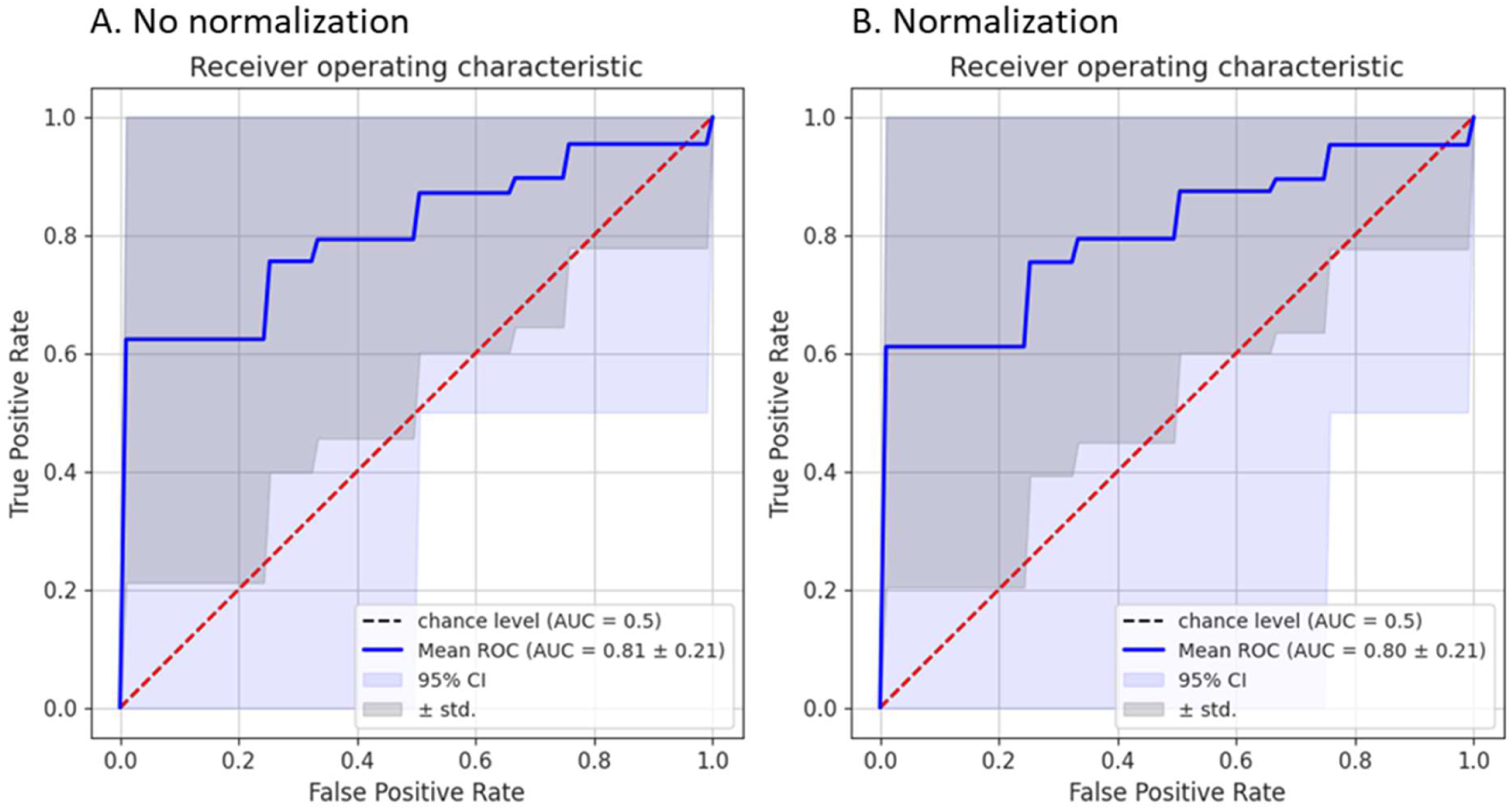
ROC curve of logistic binary regression to predict therapeutic decision using all regressors (n=54) using a 10-fold cross-validation without normalization **(A)** and with normalization **(B)**. Mean ROC curve, 95% confidence interval (CI) and standard deviation are shown.

**Table 2.** Logistic regression with selected parameters from the stepwise regression using all available regressors (n=54).

| 1. No Normalization |  |  |  |  |  |
| --- | --- | --- | --- | --- | --- |
|  | Coef. | STD.Err. | z | P > z | [0.025 0.975] |
| Bladder function subscore | -1.8436 | 0.5793 | -3.1827 | 0.0015 | -2.9789 -0.7083 |
| CHEPS T4 pathologic | 2.2958 | 0.8827 | 2.6009 | 0.0093 | 0.5657 4.0258 |
| aSCOR | 0.0428 | 0.0197 | 2.1787 | 0.0294 | 0.0043 0.0813 |
| dSEP C8 pathologic | 2.3622 | 1.1746 | 2.0110 | 0.0443 | 0.0600 4.6645 |
| 2. Normalization of automatic ratios |  |  |  |  |  |
|  | Coef. | STD.Err. | z | P > z | [0.025 0.975] |
| Bladder function subscore | -1.8436 | 0.5793 | -3.1827 | 0.0015 | -2.9789 -0.7083 |
| CHEPS T4 pathologic | 2.2958 | 0.8827 | 2.6009 | 0.0093 | 0.5657 4.0258 |
| aSCOR | 0.0428 | 0.0197 | 2.1787 | 0.0294 | 0.0043 0.0813 |
| dSEP C8 pathologic | 2.3622 | 1.1746 | 2.0110 | 0.0443 | 0.0600 4.6645 |

#### 3.4.2. Excluding the electrophysiological regressors

To gain statistical power, we also tested a model that excluded the electrophysiological regressors (missing in n=46 patients), bringing the number of patients to 100 (vs. 54 in the previous section). **Table 3** presents the logistic regressions with the significant predictors (p<0.05) from the stepwise regression without normalization (model 1) and with normalization (model 2). The resulting ROC curves are presented in Figure 5 without normalization (Figure 5a) and with normalization (Figure 5b). The model without normalization yielded an accuracy of 0.684±0.128 while the accuracy was of 0.661±0.130 with normalization. CSA ratio (model 1) and CSA ratio normalized (model 2) were a significant predictor in addition to myelopathy and the upper extremity sensory subscore.

**Figure 5.**
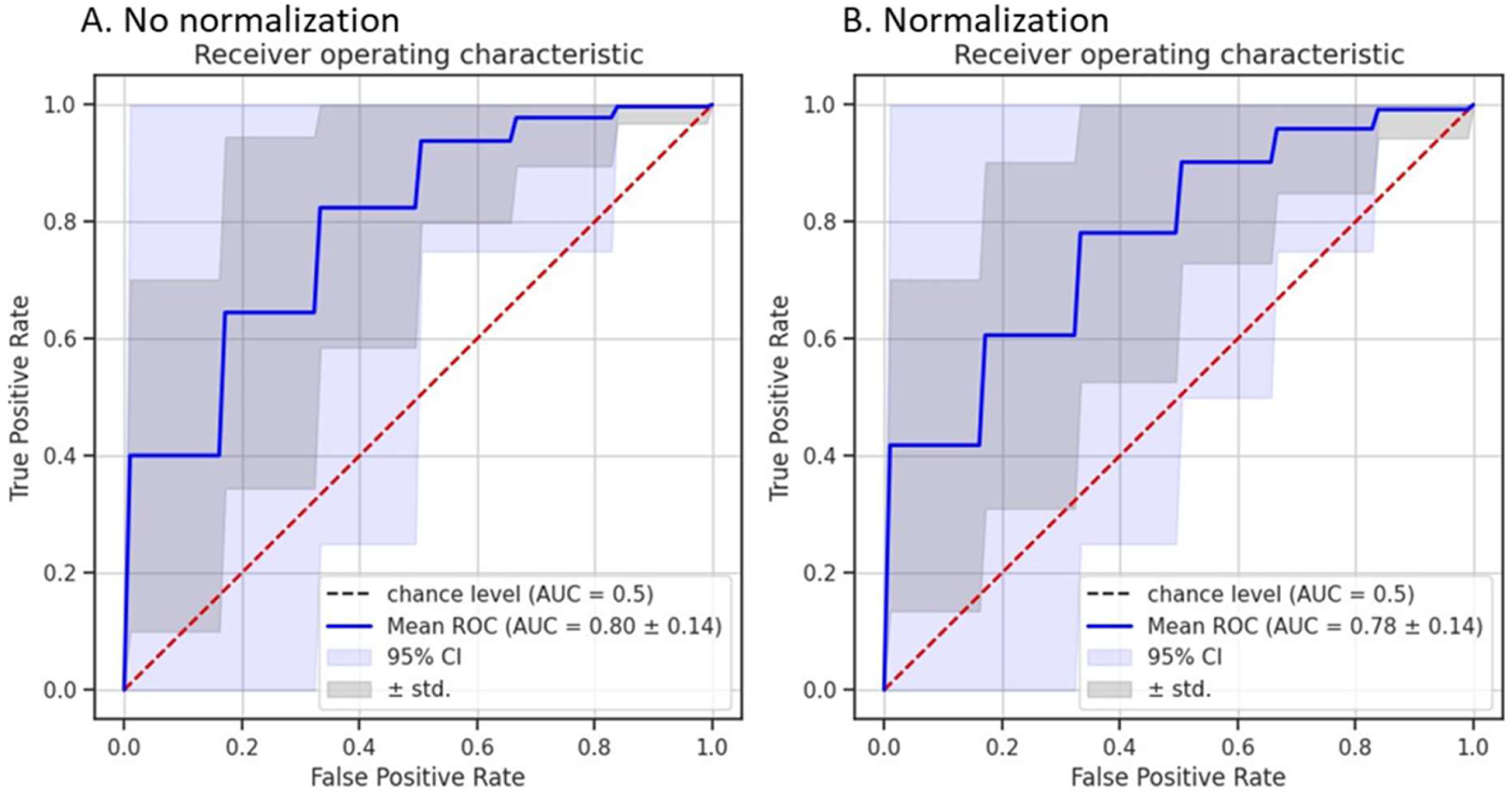
ROC curve of logistic binary regression to predict therapeutic decision without electrophysiological (n=100) using a 10-fold cross-validation without normalization **(A)** and with normalization **(B)**. Mean ROC curve, 95% confidence interval (CI) and standard deviation are shown.

**Table 3.** Logistic regression with selected parameters from the stepwise regression removing electrophysiological data (n=100).

| 1.No Normalization |  |  |  |  |  |
| --- | --- | --- | --- | --- | --- |
|  | Coef. | STD.Err. | z | P > z | [0.025 0.975] |
| **Upper extremity sensory subscore** | -0.8948 | 0.1933 | -4.6303 | <0.0001 | -1.2736 -0.5161 |
| **CSA ratio** | 0.0537 | 0.0193 | 2.7904 | 0.0053 | 0.0160 0.0915 |
| **Myelopathy** | 1.0854 | 0.5364 | 2.0233 | 0.0430 | 0.0340 2.1367 |
| 2. Normalization of automatic ratios |  |  |  |  |  |
|  | Coef. | STD.Err. | z | P > z | [0.025 0.975] |
| **Upper extremity sensory subscore** | -0.9636 | 0.2181 | -4.4176 | <0.0001 | -1.3911 -0.5361 |
| **CSA ratio normalized** | 0.0535 | 0.0215 | 2.4889 | 0.0128 | 0.0114 0.0956 |
| **Myelopathy** | 1.2102 | 0.5323 | 2.2735 | 0.0230 | 0.1669 2.2535 |

Coef.: coefficient; STD Err. standard error of coefficient; z: z value; P > z: p-value;

## 4. Discussion

We introduced an automatic method to quantify cord compression from clinical MRI scans. We extended the traditional MSCC (based on AP diameter) to other morphometrics (RL diameter, CSA, eccentricity, and solidity), and added a normalization across healthy adults to better account for variability in the spinal cord anatomy along the superior-inferior axis. We validated the proposed methodology against manual measurements and clinical scores in a cohort of 120 DCM patients and explored which metrics can predict therapeutic decision (operative/conservative). Using a stepwise binary logistic regression, we found that the upper extremity sensory subscore, CSA ratio, CSA ratio normalized and radiological signs of myelopathy were significant predictors of the therapeutic decision leading to an AUC of 80% and 78% (with normalization).

### 4.1. Correlations between morphometrics and clinical scores

The proposed MRI morphometrics significantly correlated with clinical scores (mJOA & CSA ratio: ρ=-0.29, p-value=0.004), better than the manually derived aMSCC (mJOA: ρ=0.09, p-value=0.4) and aSCOR (mJOA: ρ=-0.13, p-value=0.2). Previous studies reported no correlation between manual MRI morphometrics and mJOA^27–29^, or correlation for some morphometrics (compression ratio, MCC, and MSCC) with mJOA^12,30^. This could be caused by several factors including differences in cohort sizes and inclusion/exclusion criteria, variability in scanner field strengths (1.5T vs. 3.0T) and sequences used (axial vs. sagittal; T1w vs T2w), and intra– and inter-rater variability in manual measurements. Moreover, although mJOA is currently a widely accepted clinical score for assessing DCM patients, it has poor sensitivity, which is especially problematic in mild DCM (mJOA of 15-17)^3^.

### 4.2. Comparison of manual vs. automatic MRI measures

When assessing the relationship between the automatic and manual morphometric ratios, we found that the correlation of aMSCC with CSA ratio and CSA ratio normalized was only of ρ=-0.182 (p-value=0.047) and ρ=-0.166 (p-value=0.069), respectively (Figure 3). Given that aMSCC is calculated as the area at the maximum compressed level divided by the area at C2 vertebral level, a higher agreement with the automatic morphometric ratios was expected. Surprisingly, we found higher correlation of CSA ratio and CSA ratio normalized with aSCOR (ρ=0.395 and ρ=0.325, respectively) than with aMSCC.

In the final model (n=100), CSA ratio and CSA ratio normalized were significant predictors of the therapeutic decision while neither aSCOR or aMSCC were significant (**Table 3**).

Only a few studies have used automatic or semi-automatic measures to compute spinal cord morphometrics^5,16,17,31–33^. The majority of the studies measured the AP diameter on the mid-sagittal slice^1,12,29,34–36^. However, the mid-sagittal slice may not correspond precisely to the middle of the spinal cord due to patient scoliosis and/or sequence positioning. Additionally, manually measuring the AP diameter at the compression sites and healthy levels above/below is time consuming and prone to inter-rater variability^13,30^. Another study comparing manually vs. automatically derived morphometrics (CSA, compression ratio, AP diameter, and RL diameter) showed higher inter-rater reliability for the automatically computed morphometrics^16^.

Computing MSCC only with AP diameter does not necessarily reflect the compression severity, and might poorly quantify lateral compression^35^. Computing MSCC with other shape metrics will likely increase the ability to detect and characterise various types of compressions^37^.

Some studies directly used the AP diameter or CSA in their analysis without self-normalization with healthy levels above and below the compression sites^4,16,32,33,38^. Spinal cord morphometry has a large inter-subject variability, especially for CSA, AP and RL diameters^10,11,14^ and between males and females^11,14,15,39–41^. This suggests the need for self-normalization in cross-sectional studies.

### 4.3. Impact of across-subject normalization

We presented 2 different models predicting the therapeutic decision in a cohort of 120 DCM patients. In both models, the same automatic morphometric ratios were significant. For the analysis without electrophysiological data (n=100), the model with normalized morphometric had a slightly lower AUC, 0.78±0.14 compared to 0.80±0.14. There is no clear conclusion on the effect of this additional normalization.

The majority of DCM patients in this study had their maximum spinal cord compression located at the C5/C6 level (n=52), which aligns with previous findings^4,38^. This might reduce the overall effect of the normalization as it is designed to be beneficial especially when patients have a maximum level of compression located across different levels.

Also, the effect of the normalization might be influenced by different age ranges. The DCM cohort in this study had a mean±standard deviation age of 55.28±12.80 y.o., while the healthy control database had an age of 28.7±5.6 y.o.

The number of stenoses significantly correlated with age (ρ=0.21, p-value=0.034), suggesting a higher prevalence of compression with increasing age. Previous studies have reported that the prevalence of spinal cord compression increases with age^11,38,42^. Further work will include extending the normative database^11^ to a wider age range.

### 4.4. Predicting therapeutic decision

In the stepwise binary logistic regression, we first included demographics, automatic morphometric ratios, manual morphometric ratios, clinical data, clinical scores, and electrophysiological data. However, only 54 subjects had a complete dataset. Significant predictors for both model included bladder function subscore, CHEPS T4 pathologic, aSCOR and dSEP C8 pathologic (**Table 2**). The second analysis, excluding electrophysiological data, included 100 subjects with a complete dataset, and identified radiological signs of myelopathy (T2w hyperintensity), CSA ratio, CSA ratio normalized and the upper extremity sensory subscore guiding the decision for surgical intervention (**Table 3**). This model achieved an AUC of 80% in predicting therapeutic decisions.

This suggests that the therapeutic decision was partly based on electrophysiological assessment when available. When electrophysiological assessment was not available, the therapeutic decision was based on the anatomical MRI and radiological signs of myelopathy. When making the therapeutic decision, the neurosurgeons had access to clinical scores, anatomical MRI and electrophysiological assessment only. No morphometric ratios were provided. The therapeutic decision remains multi-factorial, which may explain the variability in the significant predictors when changing the included subjects in the analysis^28^. This suggests the need for large multi-center repositories aggregating MRI and clinical data across different demographics and clinical settings.

Hilton et al.^28^ investigated the factors in current practice that influence the decision to operate in 39 DCM patients, and concluded that only the compression ratio was a significant predictor of the decision to operate.

### 4.5. Limitations & Perspective

Our model predicting the therapeutic decision only included the morphometric ratios at the maximum level of cord compression, which may not provide a complete picture of the pathology, for instance, in multi-level stenosis. However, we did not find any association between the number of stenoses and mJOA. Additionally, MRI acquisition in a supine position may fail to accurately depict spinal cord compression associated with weight-bearing, and miss the dynamic compression visible during flexion/extension of the head^43^.

The significant morphometric ratios changed following the number of subjects, from aSCOR to CSA ratio. This could be attributed to the fact that not all types of morphometric measures necessarily reflect all the different shapes of spinal cord compression^37^.

We did not differentiate between types of surgery and did not include variables like compression ratios, or spinal canal morphometrics which previous studies found relevant for disease prognosis. Further work will include extending the automatic computation of morphometrics ratios to the spinal canal.

The computation of morphometrics directly depends on the quality of the spinal cord segmentation, which can require manual correction (45 images had to be manually corrected in this study, which took about 3 minutes per image). In cases of severe spinal cord compressions, segmenting the spinal cord becomes more challenging. The introduction of more robust segmentation models will help reduce manual intervention^44,45^. We computed the morphometrics on axial scans, with low superior-inferior resolution, potentially limiting the interpolation of the measures to the PAM50 space. Furthermore, choosing a distance from the compression site for normalization does not consider inter-subject variability of spinal cord length. Other limitations exist for the use of vertebral levels as they do not precisely estimate the spinal levels location^46^. In our pipeline, compression sites have to be manually labeled on the MRI image. The next steps will focus on the automatic detection of spinal cord compressions.

Computing morphometrics on sagittal or axial images with a large slice thickness (3-5mm) as typically acquired clinically limits the precision of the measurements considering that the spinal cord compressions only span over a few millimetres. A 3D acquisition protocol with isotropic resolution would be relevant to increase the precision of the morphometric computation as proposed in the Spine Generic protocol (T2w 0.8mm^3^)^41^.

Inter-scanner, MRI field strength and acquisition sequence variability also have to be considered. It can be minimized by using the standardized protocol^41^.

Incorporating quantitative MRI biomarkers sensitive to spinal cord tissue integrity and microstructure might also enrich the predictive model sensitivity, especially in patients with mild DCM^17,47–49^.

## 5. Conclusion

We introduced an automatic pipeline to quantify and characterize spinal cord compressions from MRI scans in DCM patients. The output metrics are normalized using a database of healthy controls. We show good agreement of the automatic morphometrics ratios with clinical scores compared to manual measures. Standardized and automatically computed cord morphometrics ensure more precision and accuracy in our ability to detect subtle cord compression due to traumatic and non-traumatic events. These objective markers have the potential to serve towards better informed therapeutic decisions.

## Data Availability

All data produced in the present study are available upon reasonable request to the authors.

## 6. Acknowledgements

We thank healthy controls and patients for participating in this study. We thank Michele Hubli, Jan Rosner, and Paulina Scheuren for providing the electrophysiological data and Carl Zipser and Martin Schubert for patient recruitment. We thank Nathan Molinier for valuable feedback on figures and fruitful discussions, Allan Martin for insightful discussions, Nick Guenther and Mathieu Guay-Paquet for their assistance with data management, and Joshua Newton and Mathieu Guay-Paquet for helping us implement the algorithm in the Spinal Cord Toolbox.

## 10. Supplementary Material

**Figure S1.**
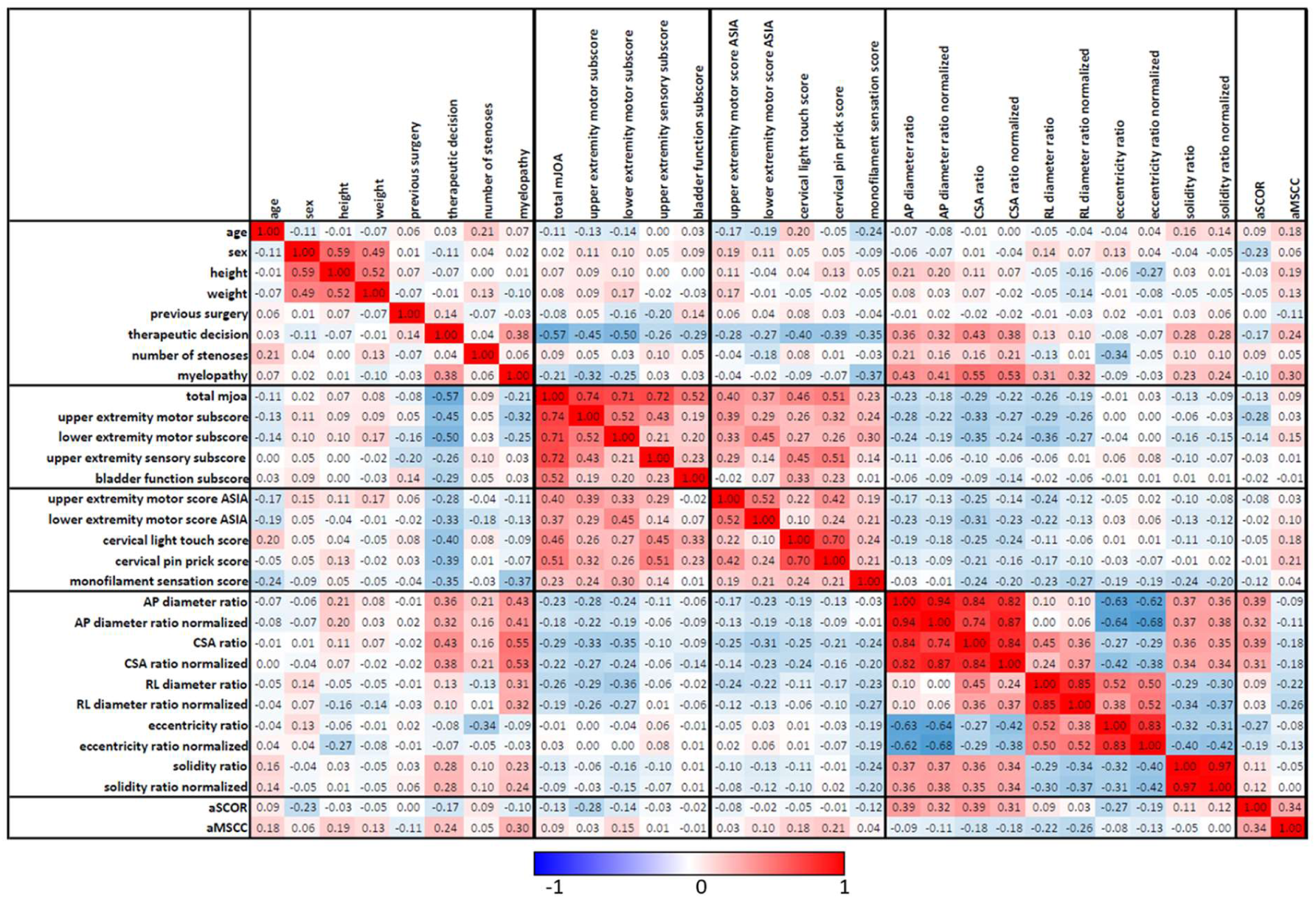
Correlation matrix. Spearman’s correlation coefficient was computed for continuous variables, phi coefficient was computed across dichotomous variables and the Point-biserial correlation was computed across continuous and dichotomous variables. Sex, previous surgery, myelopathy and the therapeutic decision were the dichotomous variables.

